# Assessing Attitudes and Perceptions of High-risk, Low-resource Communities towards Cardiopulmonary Resuscitation and Public-access Defibrillation

**DOI:** 10.1101/2024.09.26.24314460

**Authors:** Carolyn Hirsch, Bhanvi Sachdeva, Dilenny Roca-Dominguez, Jordan Foster, Kellie Bryant, Nancy Gautier-Matos, Mara Minguez, Olajide Williams, Mitchell S. V. Elkind, Shunichi Homma, Rafael Lantigua, Sachin Agarwal

## Abstract

**Background:** Layperson cardiopulmonary resuscitation (CPR) and automated external defibrillator (AED) use are vital for improving survival rates after out-of-hospital cardiac arrest (OHCA), yet their application varies by community demographics. We evaluated the concerns and factors influencing willingness to perform CPR and use AEDs among laypersons in high-risk, low-resource communities.

**Methods:** From April 2022 to March 2024, laypersons in Northern Manhattan’s Community District 12 completed surveys assessing their attitudes toward CPR and AED use before attending Hands-Only CPR training. Fisher’s Exact Test assessed differences in concerns and willingness to perform CPR and AED use across racial-ethnic groups and compared low-resource communities with high-resource groups consisting of non-clinical staff across eight ambulatory sites.

**Results:** Among 669 respondents, 64% identified as Hispanic, 58% were under 40, and 67% were female. Significant knowledge gaps were identified: 62% had never learned CPR, and 77% were unfamiliar with AEDs. Common concerns included fear of incorrect performance (67%), causing harm (56%), and legal repercussions (53%). Willingness to perform CPR was most influenced by familiarity with the victim (55% family vs. 67% non-family). The main barrier to AED use was a lack of operational knowledge (66%). Non-Hispanic Black participants expressed significantly greater concerns than their non-Hispanic White and Hispanic counterparts. Participants in high-resource settings (n=309) showed higher CPR and AED training rates with similar concerns.

**Conclusions:** Concerns regarding CPR and AED use stem from a lack of confidence and training. Targeted, culturally sensitive community interventions are essential to address these barriers, enhance preparedness, and improve OHCA survival rates.

## 1. Introduction

Despite advancements in post-cardiac arrest care, survival rates following out-of-hospital cardiac arrest (OHCA) remain alarmingly low and vary significantly by neighborhood demographics.^1,2^ For instance, New York City, with its diverse communities, reported survival rates ranging from 1.4 - 2.2%,^3,4^ starkly lower than the national average of 10-11%.^5^ These differences are also reflected in their overall rates of sustained return of spontaneous circulation (ROSC) after OHCA (25% in New York City vs. 30% nationally).^6^ A critical determinant of survival and sustained ROSC is the prompt provision of cardiopulmonary resuscitation (CPR) and automated external defibrillator (AED) by laypersons, which can substantially improve survival chances.^7,8^ However, only about 40% of OHCA victims receive immediate layperson CPR, and merely 4.6% receive defibrillation before emergency medical services arrive.^9–11^

This low rate of lifesaving interventions is influenced by various contextual factors, including geographic, socioeconomic, and demographic characteristics of both the person experiencing OHCA and the layperson. Layperson CPR rates are notably lower in socially and economically disadvantaged areas, rural regions, and among certain ethnic populations, reflecting significant geographic and demographic disparities.^12–14^ Individuals with lower socioeconomic status and Black and Hispanic individuals experience a higher incidence of OHCA^15,16^ and reduced survival.^17^ Black and Hispanic individuals are 37% less likely to receive lifesaving CPR in public spaces compared to their white counterparts.^18^

These disparities further extend to the rates of CPR training across the United States.^19^ Addressing the factors that influence a layperson’s willingness to perform CPR is essential for enhancing early resuscitation practices, improving survival rates, and reducing inequalities in high-risk, low-resource communities.

Layperson’s fear of causing harm and performing CPR incorrectly due to lack of sufficient training has been identified as a major concern and significantly reduces confidence and the likelihood of providing CPR.^14,20^ Notably, this lack of confidence has been closely linked to lower socioeconomic status and racial/ethnic composition of the community.^9,12,13,21,22^ Additional fear of legal repercussions and personal safety, including the risk of contracting infections or diseases and being in potentially unsafe environments, have collectively contributed to the hesitancy and low rates of CPR initiation, particularly in vulnerable low-resource communities.^12–14,23^

The willingness and likelihood of performing CPR during OHCA are significantly influenced by the relationship between the layperson and the individual experiencing it.^24–27^ This reluctance to assist strangers is often attributed to concerns about the unknown disease status of strangers and the potential legal consequences of performing CPR.^21^ In predominantly Hispanic/Latino neighborhoods, factors such as the mismatched age^25,28^ and sex^29–31^ of the victim compared to the layperson can decrease the likelihood of intervention. Cultural and social factors, including language barriers and a lack of community connectedness, could create reluctance to provide aid. These factors are particularly relevant in minority and economically disadvantaged neighborhoods, where such issues are more prevalent.^22,32^ These challenges underscore the need for targeted community interventions that address both relational and demographic factors to increase the rates of laypersons’ willingness to perform CPR.

A recent systematic review and meta-analysis involving 1,081,040 OHCA patients across 11 countries has highlighted the significant benefits of community-based interventions across all populations.^33^ These interventions have been associated with increased rates of layperson-initiated CPR, higher utilization of AEDs, improved survival rates, and better neurological outcomes among survivors.^33,34^ Despite these promising findings, effectively addressing barriers, concerns, and misconceptions about these interventions requires a deep understanding of community attitudes and perceptions regarding CPR and AED use.

To address this knowledge gap, the study aimed to achieve three primary objectives: First, to evaluate the knowledge and training gaps, as well as the attitudes and perceptions that may influence the willingness of laypersons in high-risk, low-resource communities in New York City to perform CPR and use AEDs. Second, to identify specific racial and ethnic differences in attitudes and perceptions to inform the development of culturally appropriate community intervention programs. Finally, to compare findings from low-resource communities with those of non-clinical staff working in high-risk, high-resource outpatient medical facilities.

## 2. Methods

### 2.1 Study Design

We used a prospective anonymous survey design to determine layperson attitudes and perceptions contributing to willingness to perform Hands-Only CPR and AED use. 3

### 2.2 Study Population

#### Low-resource, high-risk community

Between April 1, 2022, and March 31, 2024, 669 laypersons across 20 groups voluntarily participated in in-person Hands-Only CPR and AED training sessions and filled out a survey as part of a community-based initiative, Community Development-Resuscitation Education, AED, and CPR Training (CD-REACT).^35^ The groups included community-based social service organizations (45%), youth extracurricular programs (26%), parent-teacher associations (19%), and local businesses (10%).

Participants in this study were residents of Northern Manhattan’s Community District 12, which includes the neighborhoods of Washington Heights and Inwood, as well as parts of the Bronx and Harlem. With high area deprivation indices (7-10 deciles at the state level and 71-100 percentiles nationally)^36^ of these neighborhoods, they rank among the most socioeconomically disadvantaged in New York State and the country. Washington Heights, the primary recruitment area, is characterized by a high-risk, low-resource population with 68% identify as Hispanic, 20% as non-Hispanic White, and 33% with limited English proficiency.^37^ Compared to other New York City neighborhoods, Northern Manhattan residents have lower high school graduation rates, higher poverty rates (20% below the federal poverty line), and greater feelings of unsafety in their neighborhoods.^38^ Additionally, heart disease is the leading cause of death and hospitalization in this area, which ranks among the bottom 10 of 41 NYC neighborhoods for access to medical care.^38^

#### High-resource, high-risk community as a comparison group

From 10/2018 to 5/2019, the Columbia University Patient Safety Department conducted in-person training sessions covering all major outpatient medical and surgical specialties (∼1 million patient visits/year) throughout the New York Metro area. The outpatient medical sites included in this study rely on the emergency medical system for managing acute cardiac conditions requiring clinical and non-clinical staff to assume the role of first responders in providing CPR and AED administration. Approximately 80% of the non-clinical staff at eight ambulatory sites receiving training constituted the high-risk, high-resource comparison group.

### 2.3 Training

The ≈1-hour training session, in English or Spanish and identical for both low- and high-resource settings, was structured around three key components: Education, Demonstration, and Hands-on Skills Practice **(see Appendix 1** for details on the training session).

#### Pre-Training Preparation

Before the session began, participants completed a survey assessing their attitudes and perceptions of lifesaving interventions. Each participant received the American Heart Association (AHA) CPR Anytime Kit, which included a manikin for use during the training.

##### 1. Education (15 minutes)

The session commenced with an educational segment. When audio-visual equipment was available, the AHA “CPR in Action” video was shown. This video was followed by an explanation of the differences between cardiac arrest and heart attack, the benefits of Hands-Only CPR, and the Chain of Survival’s role in keeping the brain and heart of the patient alive. Participants were encouraged to share any previous experiences with administering CPR.

##### 2. Demonstration (10 minutes)

Next, a demonstration of Hands-Only CPR was conducted. This included a detailed overview of the technical aspects such as the rate, depth, quality, and duration of effective CPR.

##### 3. Hands-on Skills Practice (20 minutes)

Participants then engaged in Hands-on Skills Practice using their take-home CPR manikins. During this time, the training team circulated the room to assist with technique and answer any questions.

##### 4. AED and Q&A (15 minutes)

The final segment covered the role of an AED in the Chain of Survival, common fears associated with AED use, and typical locations where AEDs are found. The use of the AED was demonstrated through active participant engagement. The session concluded with five minutes dedicated to addressing participant questions and concerns, such as what to do if ribs are broken during CPR or the legal protections for performing CPR on a stranger.

### 2.4 Survey Instrument

We utilized a 16-item closed-question survey adapted from previous studies assessing attitudes and perceptions toward CPR/AED use in other populations.^39^ The survey, available in both English and Spanish, included demographic questions (such as age, sex, ethnicity, race, years of education, and current employment status) and inquiries about prior training in CPR and AED use. Participants were asked whether they believe cardiac arrest is the same as a heart attack, with response options of yes, no, or unsure. We also inquired about their awareness of the presence or location of an AED at their workplace or school. To gauge motivations for training, we provided several options categorized broadly into a general interest in learning, work requirements, or helping/saving a life in an emergency. Participants answered two primary questions with multiple choices regarding their general concerns about performing CPR and factors that might influence their willingness to perform CPR. They could select multiple options and add responses in an ‘Other’ category if desired. The final multiple-choice question asked for reasons they might not use an AED on a stranger who collapses in public **(see Appendix 2** for the survey).

### 2.5 Data Analysis

Descriptive statistics (percentages and proportions) were first used to report the motivations, attitudes, and perceptions associated with general willingness to perform CPR and AED use. We then used Fisher-Exact and Chi-square tests to compare the differences between community members from high-resource and low-resource settings, and the three racial-ethnic groups (Non-Hispanic White, Non-Hispanic Black, and Hispanic) for general concerns and factors associated with willingness to perform CPR. P-value <0.05 was considered statistically significant. Data was compiled and analyzed using STATA 18.0.

## 3. Results

### 3.1 Low-resource, high-risk community

#### 3.1.1 Demographics

Of the 669 laypersons who participated, 58% were under 40 years old. The majority identified as female (67%, n=445) and Hispanic (64%, n=426). Additionally, 24% (n=163) identified as Black and 56% (n=373) as White. Of the 163 Black participants, 46% (n=75) were non-Hispanic, while 23% (n=84) of the 373 White participants identified as non-Hispanic. While most participants (66%) had received high school or more education, only 41% were employed at the time of the survey.

#### 3.1.2 Knowledge gaps regarding CPR

Most participants (65%) had either never learned CPR or were unsure about it, and over half (55%) responded either incorrectly or were unsure about cardiac arrest being the same as a heart attack **(Table 1)**.

**Table 1.** Demographics and Characteristics of Laypersons in Low-Resource Communities.

| **Participants Characteristics (N=669)** | **% (n)** |
| --- | --- |
| Age |  |
| <18 | 26 (170) |
| 18-40 | 33 (216) |
| 41-60 | 23 (157) |
| 61-80 | 16 (103) |
| >81 | 2 (16) |
| Sex |  |
| Male | 31 (206) |
| Female | 67 (445) |
| Other | 1 (4) |
| Decline to respond | 1 (5) |
| Race |  |
| American Indian | 5 (34) |
| Asian/Pacific Islander | 12 (77) |
| Black | 24 (163) |
| White | 56 (373) |
| Decline to respond | 3 (22) |
| Ethnicity |  |
| Hispanic/Latino | 64 (426) |
| Non-Hispanic/Latino | 32 (217) |
| Decline to respond | 4 (26) |
| Education Status |  |
| <High school | 29 (188) |
| >High school | 66 (425) |
| Decline to respond | 5 (30) |
| Occupational Status |  |
| Employed | 41 (270) |
| Unemployed | 16 (106) |
| Student | 27 (175) |
| Homemaker | 1 (10) |
| Retired | 15 (97) |
| Have you ever learned CPR before? |  |
| Yes | 35 (230) |
| No | 62 (409) |
| Unsure | 3 (22) |
| Is Cardiac Arrest the same as a Heart Attack? |  |
| Yes | 22 (146) |
| No | 45 (292) |
| Unsure | 33 (212) |
| Have you ever learned how to administer an AED? |  |
| • Yes | 17 (117) |
| • No | 77 (504) |
| • Unsure | 6 (37) |
| Do you know if there is an AED in the building where you work or study? |  |
| • I am certain there is an AED and know where it's located | 22 (136) |
| • I am certain there is no AED | 16 (102) |
| • I'm not sure if there is an AED | 62 (386) |

#### 3.1.2 Motivation for training

When asked about their primary motivation for taking the course, the most frequently reported response was the desire to help save a life in an emergency, with 55% of participants (n=361) citing this reason. This motivation was followed by a general interest in learning, reported by 43% of participants (n=287). Other notable motivations included the ability to assist a child or an ill family member (18%; n=122) and a work requirement (13%; n=86) (**Figure 1**).

**Figure 1.**
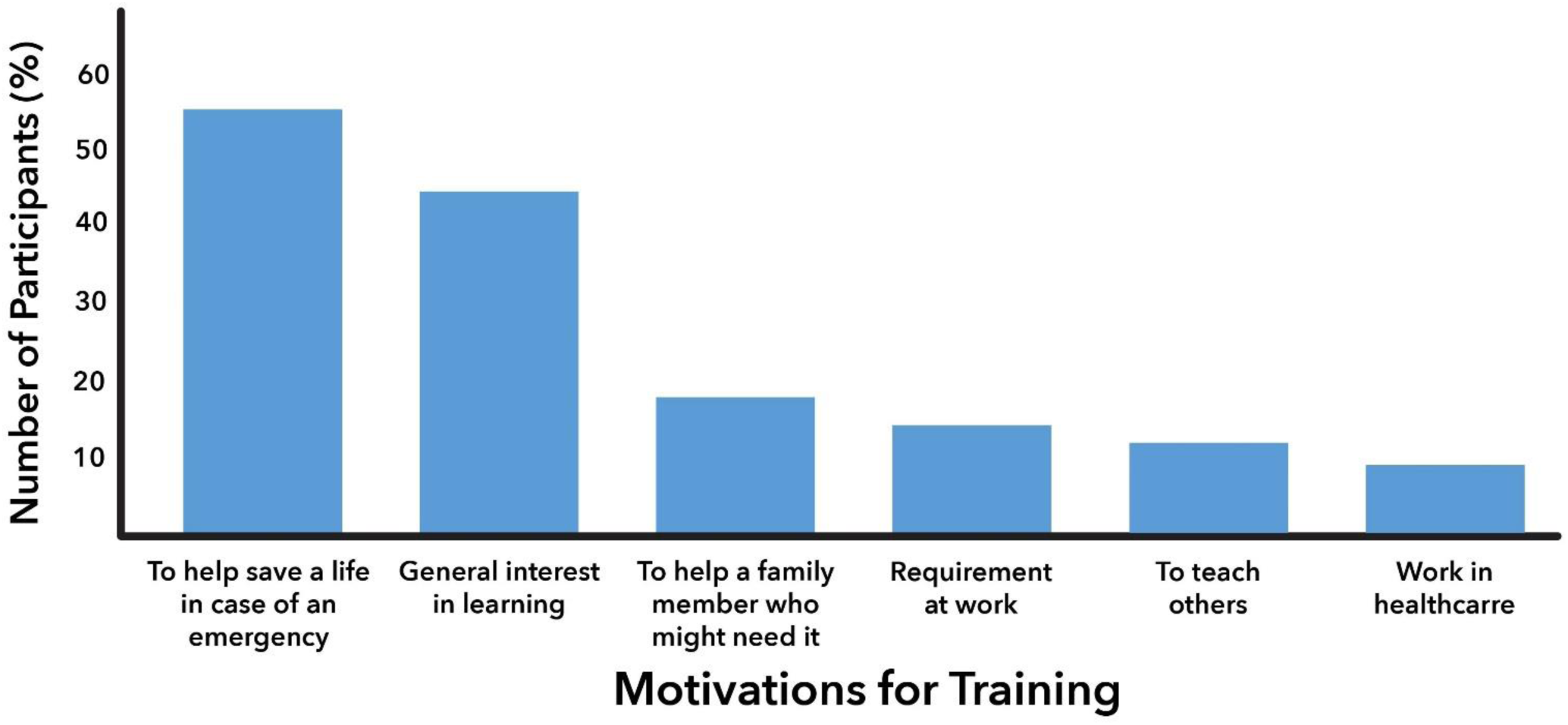
Motivations for CPR/AED Training Participation Among Laypersons in Low-Resource Communities

#### 3.1.3 General concerns with performing CPR

The primary concern among participants was the fear of performing CPR incorrectly due to insufficient training (67%, n=423). This was followed by concerns about potentially harming the person experiencing OHCA (56%, n=353). A close third concern was the fear of legal repercussions or lawsuits, particularly if the person were to die, reported in 53% of responses (n=336) (**Table 2**).

**Table 2.** Concerns About Performing CPR in Low-Resource Communities: A Comparison Across Racial-Ethnicities and with High-Resource Communities.

| Concerns | Low-resource Communities (N=632) | Hispanic (N=402) | Non-Hispanic Black (N=72) | Non-Hispanic White (N=80) | p-value | High-resource Communities (N=292) | p-value |
| --- | --- | --- | --- | --- | --- | --- | --- |
|  | % (n) | % (n) | % (n) | % (n) |  | % (n) |  |
| Performing CPR incorrectly | 67 (423) | 64 (256) | 79 (57) | 64 (51) | 0.03 | 63 (184) | 0.3 |
| Fear of harming the victim | 56 (353) | 49 (198) | 75 (54) | 61 (49) | <0.01 | --- |  |
| Afraid of getting sued if the person dies | 53 (336) | 53 (211) | 61 (44) | 55 (44) | 0.4 | 47 (137) | 0.2 |
| Catching a disease | 30 (212) | 30 (120) | 49 (35) | 44 (35) | 0.01 | 32 (92) | 0.7 |
| Being wrongfully accused of sexual harassment | 24 (153) | 23 (93) | 17 (12) | 23 (18) | 0.5 | --- |  |
| Somebody else would do it better | 29 (184) | 23 (94) | 51 (37) | 28 (22) | <0.01 | 23 (67) | 0.3 |
| Hurting myself while I provide CPR | 13 (82) | 12 (50) | 15 (11) | 14 (11) | 0.7 | 12 (35) | 0.9 |
| Other | 6 (37) | 7 (30) | 1 (1) | 3 (2) | 0.06 | 3 (8) | 0.7 |

#### 3.1.4 Factors influencing willingness to perform CPR

The top two factors influencing the willingness to perform CPR were in relation to the familiarity with the person experiencing OHCA. Participants expressed more willingness to perform CPR on family members (55%, n=325) or on someone they knew (67%, n=398). The third most important factor was whether the layperson was a medical professional or not, reported in 34% of responses (n=198). Among demographic characteristics, age was considered more frequently (34%, n=203) compared to sex (20%, n=120) or race/ethnicity (14%, n=81) of the person experiencing OHCA **(Table 3).**

**Table 3.** Factors Associated with Willingness to Perform CPR in Low-Resource Communities: A Comparison Across Racial-Ethnicities and with High-Resource Communities.

| Willingness Factors | Low-resource Communities (N=590) | Hispanic 65% (N=369) | Non-Hispanic Black (N=68) | Non-Hispanic White (N=76) | P-value | High-resource Communities (N=243) | P-value |
| --- | --- | --- | --- | --- | --- | --- | --- |
|  | % (n) | % (n) | % (n) |  |  | % (n) |  |
| Knowing the person, other than family | 67 (398) | 63 (233) | 82 (56) | 70 (53) | <0.01 | 48 (116) | <0.01 |
| Family member | 55 (325) | 56 (205) | 57 (39) | 54 (41) | 0.9 | 39 (95) | <0.01 |
| Medical professional | 34 (198) | 30 (111) | 41 (28) | 39 (30) | 0.08 | 39 (95) | 0.4 |
| Age of patient | 34 (203) | 35 (130) | 32 (22) | 33 (25) | 0.8 | 21 (51) | 0.07 |
| The responder has a physical disability | 25 (147) | 24 (89) | 23 (16) | 26 (20) | 0.8 | 19 (45) | 0.4 |
| Gender of patient | 20 (120) | 21 (76) | 16 (11) | 20 (15) | 0.7 | 10 (24) | 0.2 |
| Race/ethnicity of patient | 14 (81) | 12 (46) | 22 (15) | 7 (5) | 0.03 | 8 (19) | 0.5 |
| Other | 8 (46) | 8 (28) | 3 (2) | 9 (7) | 0.3 | 19 (47) | 0.1 |

#### 3.1.5 Knowledge gaps and barriers associated with the use of public access defibrillators

Significant learning gaps were identified with 77% (n=504) of participants reporting that they had never learned how to use an AED and an additional 6% (n=37) being unsure. A majority (62%, n=386) were unaware of whether an AED was available at their workplace or school, while 16% (n=102) were certain that no AED was present **(Table 1)**.

The primary barrier to public access defibrillation was a lack of knowledge on how to use AEDs (66%, n=417). Additionally, concerns about causing harm to the individual experiencing OHCA (25%, n=158) and perceptions of difficulty in using an AED (17%, n=108) were also commonly cited factors **(Table 4)**.

**Table 4.** Barriers Associated with Public Access Defibrillation in Low-Resource Communities: A Comparison Across Racial-Ethnicities.

| Barriers | Low-resource Communities (N=635) | Hispanic 67% (N=408) | Non-Hispanic Black (N=66) | Non-Hispanic White (N=80) | P-value |
| --- | --- | --- | --- | --- | --- |
|  | % (n) | % (n) | % (n) | % (n) |  |
| Don't know how to use | 66 (417) | 67 (273) | 59 (39) | 64 (51) | 0.6 |
| Will harm the other person | 25 (158) | 22 (90) | 41 (27) | 25 (20) | <0.01 |
| Difficult to Use | 17 (108) | 13 (55) | 29 (19) | 20 (16) | <0.01 |
| Think I am not allowed to use | 15 (94) | 14 (59) | 28 (19) | 8 (6) | <0.01 |
| Will not help the person | 4 (26) | 4 (16) | 9 (6) | 1 (1) | 0.06 |

#### 3.1.6 Stratified analysis by race-ethnicities

##### 3.1.6.1 Knowledge gaps

Hispanic participants reported the lowest rate of previous CPR training (29%, n=121), followed by non-Hispanic Blacks (50%, n=38) and non-Hispanic Whites (63%, n=53; p<0.01). Similar disparities were seen in AED training, with the lowest rates among Hispanic (13%, n=55) and non-Hispanic Black participants (24%, n=18), compared to non-Hispanic Whites (39%, n=33; p<0.01).

Regarding whether cardiac arrest is the same as a heart attack, the majority of Hispanic (52%, n=215) and non-Hispanic Black (63%, n=46) participants were either incorrect or unsure, compared to 53% (n=42; p=0.09) of non-Hispanic Whites.

The ranking of the top three concerns, factors affecting the willingness to provide CPR, and barriers related to AED use were consistent across all racial-ethnic groups. However, non-Hispanic Black participants expressed significantly higher concerns than non-Hispanic White and Hispanic participants. Specifically, 79% of non-Hispanic Black participants worried about performing CPR incorrectly (64% for both other groups, p=0.03). Additionally, 75% feared harming the person in cardiac arrest (61% and 49%, p<0.01), and 49% were concerned about contracting a disease (44% and 30%, p=0.01). Lastly, 51% believed someone else would perform CPR better compared to 28% and 23% in the other groups (p<0.01) **(Table 2)**.

Non-Hispanic Black participants were significantly more likely than non-Hispanic White and Hispanic participants to indicate that certain factors influence the willingness to perform CPR. Notably, 82% of non-Hispanic Black participants reported that knowing a person in some capacity (not including family members) affects their desire to act, compared to 70% of non-Hispanic White and 63% of Hispanic participants (p<0.01). There were no differences found across three groups in willingness to perform CPR on a family member (57% vs 54% vs 56%, p=0.9). Additionally, 22% of non-Hispanic Black participants felt that the race-ethnicity of the person experiencing cardiac arrest would impact someone’s willingness to act, compared to 7% and 12% in the other groups (p=0.03) **(Table 3)**.

A significantly higher percentage of non-Hispanic Black participants expressed barriers to using an AED compared to non-Hispanic White and Hispanic participants. Most significant barriers included a belief that using an AED could harm the person in cardiac arrest (41% vs 25% vs 22%, p<0.01), difficulty using an AED (29% vs 20% vs 13%, p<0.01), or a feeling they were not allowed to use one (28% vs 8% vs 14%, p<0.01) **(Table 4)**.

### 3.2 Characterizing the high-resource community and comparisons with low-resource community

#### 3.2.1 Demographics and knowledge gaps

Compared to participants from low-resource communities, non-clinical staff members from outpatient sites constituting the high-resource comparison group (n=309) had significantly higher percentages of females (79% vs. 67%, p<0.01) and a lower percentage of individuals of Hispanic ethnicity (32% vs. 64%, p<0.01). Additionally, a significantly greater percentage of the high-resource group had previously learned CPR (60% vs. 38%, p<0.01) and the use of an AED (32% vs. 17%, p<0.01).

#### 3.2.2 Motivation for training

Similar to participants from low-resource communities, the primary motivation for training among the participants in the high-resource group was a desire to help save a life in an emergency (43%, n=114), followed by a general interest in learning (37%, n=98).

#### 3.2.3 General concerns with performing CPR

High-resource group, similar to their low-resource community members, cited performing CPR incorrectly (63%, n=184) and the fear of legal repercussions (47%, n=137) as their top concerns. There were no statistically significant differences between the two groups regarding any of the concerns **(Table 2)**.

#### 3.2.4 General willingness to perform CPR

Although significantly lower than among participants from low-resource communities, knowing the patient in any capacity other than family (48% vs. 67%, p<0.01) and whether the person was a family member (39% vs. 55%, p<0.01) were still the top two factors identified among the high-resource group **(Table 3)**.

## 4. Discussion

Our study identified significant gaps in CPR-related knowledge and training, as well as highlighted critical concerns and factors influencing laypersons’ willingness to perform CPR and use AEDs, particularly in high-risk, low-resource, predominantly Hispanic communities.

The majority of laypersons, who were predominantly young, female, and Hispanic, demonstrated significant gaps in CPR and AED knowledge. Specifically, 62% had never learned CPR, and 77% had not received AED training. These findings highlight a pressing need for increased educational efforts and resources. Despite more than half expressing helping save a life as their primary motivation, concerns including fear of performing CPR incorrectly, causing harm, and potential legal repercussions were highly prevalent. This finding is consistent with previous research^1,14,20,40,41^ and is irrespective of CPR training status. Addressing these specific fears through improved educational materials as part of CPR training and including them in public awareness campaigns, such as those related to New York State’s Good Samaritan Law^42^ and the federal Cardiac Arrest Survival Act,^43^ will help these communities cultivate a culture of empowerment.

Our study also found that familiarity with the patient—whether familial or otherwise—significantly influenced the willingness to perform CPR. This aligns with other studies^1,34^ reporting a higher willingness to assist family members compared to strangers. Acknowledging this aspect of ‘human nature’ is essential for informing strategies such as engagement activities (e.g., workshops, social gatherings, volunteer days) that foster interaction among community members. Creating platforms for sharing experiences and partnering with local organizations—such as schools, nonprofits, and businesses—can broaden outreach and resources, reinforcing a sense of community. Although not as commonly reported by our participants, age-,^13^ and sex-based^31^ disparities in willingness to perform CPR have also been identified in other studies.

### 4.1 Disparities and cultural considerations

There are known geographic and ethnic disparities in CPR use. Hispanic and Black neighborhoods receive significantly lower rates of bystander CPR compared to predominantly White neighborhoods.^1,44^ Qualitative studies have indicated that economically disadvantaged Black and Hispanic communities often experience reduced community connectedness, which can make individuals less likely to assist strangers. In our study, non-Hispanic Black participants were significantly more concerned about incorrectly performing CPR, diffusion of responsibility, causing harm, or contracting a disease, compared to their non-Hispanic White and Hispanic counterparts. This group also found AED use to be more challenging.

Despite having the lowest rates of CPR and AED training and the highest knowledge deficits, Hispanic participants appeared to be the most willing to perform CPR. This may be attributed to the unique characteristics of Northern Manhattan, a densely populated and closely-knit community with a high proportion of foreign-born residents (48%). In this area, Dominicans represent the largest group (62%), followed by Mexicans (10.5%) and Puerto Ricans (approximately 7%).^37^ The strong community ties and cultural cohesion within these groups, combined with enhanced training opportunities, could empower individuals and increase their willingness to perform CPR and use AEDs. Tailoring interventions to address specific concerns and leveraging cultural strengths has been shown to improve the effectiveness of community-based programs.^33^ This is particularly important for high-risk neighborhoods, where they are also less likely to be trained in CPR.^45,46^

### 4.2 Comparison with high-resource group

Unsurprisingly, non-clinical staff members from high-resource outpatient sites had higher rates of prior CPR and AED training. Despite this, the concerns about performing CPR incorrectly and legal consequences mirrored those of the low-resource lay participants, suggesting that such concerns are pervasive across different community settings. Previous research shows that outpatient professionals, who are often in situations requiring CPR, feel underqualified or undertrained.^47^ Studies comparing outcomes of OHCA in clinics versus other public locations have found no significant differences in bystander CPR rates, ROSC, hospital admissions, or survival to discharge.^48^ Given that such emergencies are frequent in high-risk outpatient settings and a dedicated code team is absent, ongoing reinforcement through refresher courses and hands-on practice is essential to enhance both comfort and competency.

### 4.3 Implications for Community Interventions

Our data underscore the importance of designing culturally sensitive and contextually relevant interventions to improve CPR and AED training. In low-resource communities, where training gaps and high levels of concern are prevalent, efforts should focus on reducing barriers, building confidence, and enhancing access to training resources. Additionally, combining community-based and health system interventions, including CPR and AED training, as well as dispatcher-assisted CPR, could have a synergistic effect.^33^ Mass media campaigns and awareness initiatives may further contribute to a positive shift in attitudes towards resuscitation and increase bystander willingness to perform CPR.^49^

### 4.4 Strengths

The study was part of a Northern Manhattan initiative that illustrates how collaboration between healthcare institutions and community stakeholders can enhance public health outcomes. A key aspect of the project involved partnerships among various departments and schools within an academic medical center, including the schools of medicine, nursing, and public health, as well as a hospital and community organizations, including inner-city middle and high schools. Clinicians, public health researchers, and trainees served as health educators, extending their roles beyond clinical and research settings. Training sessions were held at community partner sites, such as schools and organizational headquarters, to effectively meet communities where they are. These partnerships ensure that community members have access to relevant knowledge and tools for responding to medical emergencies, ultimately improving the overall health and well-being of the population we serve. To the best of our knowledge, this is the only assessment of lay communities’ attitudes and perceptions conducted in the post-COVID-19 pandemic era.

### 4.5 Limitations and Future Research

This study has several limitations. The cross-sectional design limits our ability to assess changes in CPR knowledge and attitudes over time, particularly post-training. Our sample consisted of highly motivated participants, which may not be representative of the general population. Although our sample mirrored the racial-ethnic makeup of the study neighborhood,^50^ there was a lower proportion of non-Hispanic Black and non-Hispanic Whites available to make meaningful interpretations and could only be considered as hypothesis-generating evidence. Additionally, the reliance on self-reported data may introduce biases.

Future research should involve longitudinal studies to evaluate the long-term impact of interventions and assess the effectiveness of different training methods. Further research could also explore how social and cultural factors shape attitudes toward CPR and AED use across diverse populations. Early and continuous training, starting with middle and high school students, could help build a culture of action and prepare future laypersons to act promptly in cardiac emergencies.^51^

### 4.6 Conclusion

Our study highlights significant gaps in knowledge and training on immediate resuscitation measures following OHCA and characterizes laypersons’ attitudes toward CPR and AED use across various community settings. Marginalized racial-ethnic groups often express greater and more specific concerns. Targeted, culturally sensitive interventions can address these disparities, improving laypersons’ preparedness and enhancing survival rates for OHCA. By understanding and mitigating community-specific barriers, we can foster a more inclusive and effective approach to emergency cardiovascular care.

## Data Availability

Data will be available on request.

## Funding

Sachin Agarwal receives support from the National Heart Lung and Blood Institute (R01HL153311) and the National Institute of Neurological Disorders and Stroke (R01NS127959). Any-time CPR kits given to the participants were donated by the American Heart Association.

## Conflicts of Interest

All authors have disclosed that they do not have any potential conflicts of interest.

## Compliance with all instructions to authors

This manuscript has complied with all instructions given to authors.

## Authorship requirements met

All authors have made substantial contributions to the following as outlined:

1. substantial contributions to conception and design, acquisition of data, or analysis and interpretation of data,
2. drafting the article or revising it critically for important intellectual content,
3. final approval of the version to be published,
4. agreement to be accountable for all aspects of the work in ensuring that questions related to the accuracy or integrity of any part of the work are appropriately investigated and resolved.

The final manuscript has been approved by all authors.

## Manuscript not published elsewhere

There is no overlap with previous publications, and I confirm that the manuscript, including related data, figures, and tables, has not been published previously and that the manuscript is not under consideration elsewhere.

## Appendix 1. Community Development-Resuscitation Education, AED, and CPR Training (CD-REACT) Trainer Guide

There are 3 main elements of the **≈1-hour training: Education**, ***Demonstration***, ***Hands-on Skills Practice.*** Before the training: *the coordinators from community affairs handle setup. Participants fill out a survey on their knowledge and beliefs regarding lifesaving interventions and receive a <u>CPR Anytime Kit</u> with Mannikin*.

1. <u>Education (15 minutes)</u>

- **Introduce yourself** to the audience and share what skills the audience will learn today.

- If AV is available, we will play the American Heart Association’s <u>CPR in Action video</u>.
- **Ask the audience:** What is <u>cardiac arrest vs. heart attack</u>? Do you have experience with giving CPR?
- **Top lines:** Hands-Only CPR can triple the chance of survival. <u>Chain of survival</u>: identify cardiac arrest ◊call 911◊start CPR, and administer Automatic External Defibrillator (AED) if available, while you wait for paramedics. <u>Goal:</u> keep the brain alive; CPR pumps blood to the brain when the heart cannot.
2. <u>Demonstration I: Hands-Only CPR (15 minutes)</u>

- **Set the scene**: You see a person pass out in a grocery store. **Ask the audience:** What do you do first?

- Confirm unresponsiveness: Ask the person loudly, “Are you okay?”-NO RESPONSE
- Check the breathing: if any abnormality in the breathing pattern → Call 911
- Do <u>not</u> attempt to check the pulse to confirm – it is unreliable; you could be feeling your own pulse.
- **Demonstrate CPR** and emphasize important elements:

- Place your dominant hand on top of the non-dominant hand touching the chest and interlock fingers.
- Find the area just below the center of the chest above the diaphragm. Keep your elbows locked and bend forward so your body weight can help you with compressions
- 100 compressions per minute, two inches deep, allow the chest to come back to its normal state, uninterrupted for two minutes
- NO more mouth-to-mouth breathing (not beneficial in adults). Avoid all CPR interruptions. The AHA still recommends CPR with compressions and breaths for infants, children, victims of drowning or drug overdose, and people who collapse due to breathing problems.
3. <u>Hands-on Skills Practice (set to music) (20 minutes)</u> Participants practice on their take-home CPR manikins, and the training team circles the room to help with form and answer questions. The manikins make a clicking noise when CPR is administered correctly.
4. <u>Demonstration II: AED use (10 minutes):</u> *Return to the front of the room*.

- **Ask the audience:** Where can you find an AED? Airports, schools, restaurants, libraries, gyms. Do you know where an AED is in your workplace/school? If I open this device right now, will it shock me?
- Ask a **volunteer** to come to the front & demonstrate AED use: place pads, provide shock, and resume CPR.
5. <u>Call for a volunteer to run the ‘Chain of Survival’ (described above) and conclude (5 minutes).</u> **FAQs:**

- <u>What if I break someone’s ribs?</u> Broken ribs can be repaired. You are saving a life.
- <u>Can you get sued for giving CPR to a stranger?</u> No, <u>NYS Good Samaritan Law</u> protects you.
- <u>Is this training official CPR certification?</u> No, but you will receive a Certificate of Completion.

## Appendix 2. Community Development Resuscitation Education And CPR/AED Training (CD-REACT)

### Anonymous Brief Awareness Questionnaire

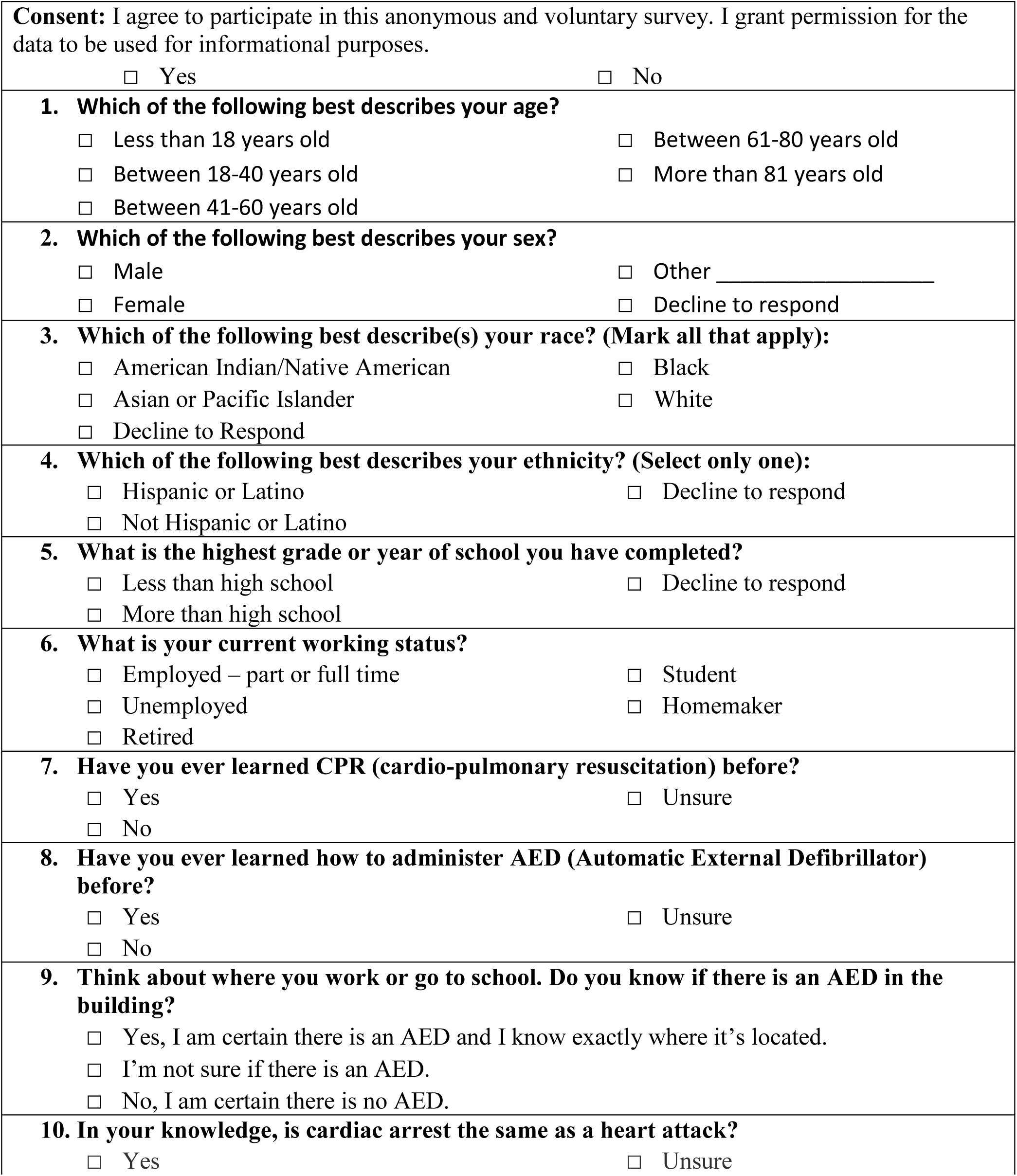
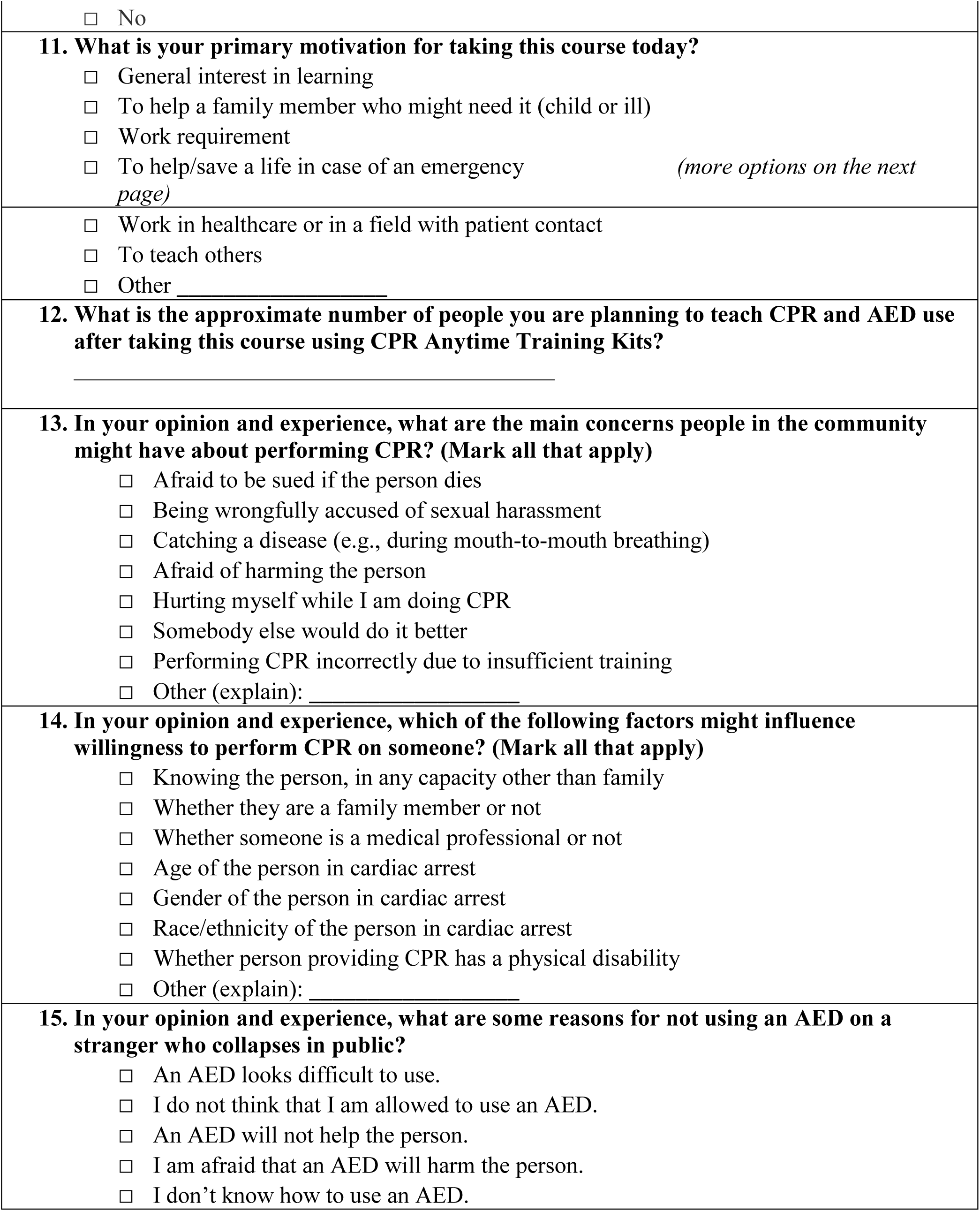

## Notes

### Competing Interest Statement

The authors have declared no competing interest.

### Author Declarations

The Columbia University Institutional Review Board approved the study protocol (IRB-AAAR8497).

